# Automated transcription in primary progressive aphasia: Accuracy and effects on classification

**DOI:** 10.64898/2026.02.24.26346981

**Authors:** Natasha Clarke, Brittany Morin, Christophe Bedetti, Rian Bogley, Sophie Pellerin, Bérengère Houzé, Siddarth Ramkrishnan, Zoe Ezzes, Zachary A. Miller, Maria Luisa Gorno-Tempini, Jet M.J. Vonk, Simona Maria Brambati

## Abstract

**INTRODUCTION:** Connected speech analyses can help characterize linguistic impairments in primary progressive aphasia (PPA) and classify variants, however, manual transcription of speech samples is time-consuming and expensive. Automated speech recognition (ASR) may be efficacious for transcribing PPA speech.

**METHODS:** Transcripts of picture descriptions (109 PPA, 32 healthy controls (HC)) were generated using a manual, automated (Whisper) or semi-automated approach including a quality control (QC) step. We evaluated transcript accuracy, the reliability of ASR-derived linguistic features, and classification performance.

**RESULTS:** Whisper demonstrated lowest error rates for HC, followed by semantic, logopenic and non-fluent PPA variants. Errors correlated with overall disease severity for semantic and logopenic variants. QC of Whisper outputs reduced errors and improved the reliability of linguistic features. Overall, ASR-derived features achieved better classification performance than manual transcription features.

**DISCUSSION:** Results support the use of off-the-shelf ASR for scalable, cost-efficient transcription of PPA speech and classification.

## Background

Primary progressive aphasia (PPA) is a clinical syndrome characterized by the gradual and relatively selective decline of speech and language abilities, caused by progressive neurodegeneration of left-hemisphere language networks^1,2^. Three main PPA variants are recognized. The non-fluent/agrammatic variant (nfvPPA) is characterized by effortful, halting speech caused by apraxia of speech and/or agrammatism. The semantic variant (svPPA) involves fluent but empty speech, with marked impairments in naming and single-word comprehension reflecting a loss of semantic knowledge, while, in contrast, the logopenic variant (lvPPA) is marked by word-finding difficulties, impaired sentence repetition, and phonological errors^1,3,4^. Identification and accurate subtyping of PPA is critical for clinical management and research, yet it remains challenging due to symptom overlap and variability in presentation across individuals.

Connected speech (CS) tasks, such as describing a picture, are used widely in research and clinical settings to assess PPA speech. Rich linguistic features (e.g., lexical diversity, syntactic complexity and fluency) can be automatically extracted from CS samples quickly, objectively and at low-cost^5^, and input to machine learning models to classify PPA patients with high accuracy^6–8^, showing promise for clinical adoption in diagnosis and monitoring. However, the speech-to-text transcription process remains a challenge. Manual transcription is considered the gold standard, but is time-consuming and expensive, and can also be subject to human error, as well as inter-rater and intra-rater variability. Automated speech recognition (ASR) systems, which use AI to convert spoken words to text, may be a fast, low-cost alternative to manual transcription.

ASR transcription performance, assessed using the word error rate (WER), is accurate for younger and middle-aged adults^9^, but has historically shown lower accuracies for older adults, who exhibit changes in voice quality^10^, and pathological speech, since systems lack the large amounts of annotated data needed to help train recognition of speech errors^11^. Early PPA studies reported high WERs, ranging from 61% - 67.7% and 37% - 73.1% for nfvPPA and svPPA respectively^12^. Despite these low accuracies, downstream machine learning models using ASR-derived features for classification achieved strong performance^13^, even out-performing features derived from manual transcription^12^, indicating robustness of classifiers to ASR error^14^.

Off-the-shelf ASR systems have significantly advanced in recent years, with access to larger datasets, large language models and sophisticated transformer-based architectures. Tools such as Google’s Speech-to-Text^15^ and OpenAI’s Whisper^16^ offer accessible, freely available application programming interfaces (APIs), enabling broader adoption in research and clinical settings, with the potential to improve reproducibility across labs. In Alzheimer’s disease, use of such systems followed by linguistic feature extraction and classification has demonstrated strong performance^17,18^, additionally demonstrating that errors in transcription may actually contain clinically meaningful information that can be used by the classifier to improve performance^19^. However, the accuracy of such systems on PPA speech, and the impact on machine-learning based classification, has not been tested. If comparable to manual transcription, off-the-shelf ASR systems could ensure scalable, cost-efficient CS analyses.

We tested the efficacy of AI-powered automated transcription for PPA variants using OpenAI’s Whisper^16^. Compared to other ASR systems, Whisper is less sensitive to speaker age and accents, tends to perform well on a variety of datasets^20^, and it runs locally, making it well-suited for processing confidential clinical data. Using manual transcription as the ground-truth, we assessed transcription accuracy, the reliability of linguistic feature values derived from ASR-transcriptions, and classification performance using features as input. We focused on classification between PPA variants and healthy controls, and between fluent PPA variants, since overlapping symptoms between lvPPA and svPPA can make the distinction more challenging. Classification involving nfvPPA was restricted to comparison with healthy controls, as our primary goal was to assess ASR performance rather than to optimise clinical subtype discrimination. We hypothesised that svPPA and lvPPA transcriptions would contain few errors, and that ASR-derived features would perform equally well in machine learning classifications as features derived from manual transcripts. We also hypothesised that the motor speech impairments in nfvPPA would impact transcript quality, resulting in more errors and lower performance in machine learning classifications for this variant.

## Methods

### Participants

This study included 151 participants from the University of California, San Francisco (UCSF) Memory and Aging Center (MAC): 39 diagnosed with svPPA, 40 with lvPPA, 40 with nfvPPA and 32 cognitively healthy controls (HC) (see Table 1). Participants with PPA were selected from the UCSF MAC research cohort, which includes individuals referred for evaluation of cognitive, language, or behavioral symptoms and who undergo multidisciplinary assessments, including neurological exams, neuropsychological testing, and structural imaging. HC were primarily drawn from the Brain Aging Network for Cognitive Health (BrANCH) and were screened to rule out neurological and psychiatric conditions. Figures showing the distribution of participant demographics can be found in the supplementary materials.

**Table 1.** Participant demographics. Values are expressed as mean ± std (range).

| Diagnosis | N | N Female | Age | Education | MMSE | CDR Sum of boxes |
| --- | --- | --- | --- | --- | --- | --- |
| HC | 32 | 20<br>(63%) | 70.78 ± 7.03<br>(53–87) | 17.19 ± 2.04<br>(12.0–20.0) | 29.33 ± 0.76<br>(28.0–30.0) | 0.0 ± 0.0<br>(0.0–0.0) |
| lvPPA | 40 | 22<br>(55%) | 62.95 ± 7.52<br>(51–81) | 16.62 ± 2.86<br>(12.0–24.0) | 19.75 ± 7.54<br>(3.0–30.0) | 3.58 ± 2.54<br>(0.0–11.0) |
| nvPPA | 40 | 25<br>(63%) | 66.88 ± 7.84<br>(51–83) | 15.95 ± 3.25<br>(12.0–28.0) | 26.78 ± 3.28<br>(12.0–30.0) | 1.49 ± 1.49<br>(0.0–5.0) |
| svPPA | 39 | 18<br>(46%) | 63.38 ± 7.28<br>(50–88) | 16.56 ± 2.11<br>(12.0–21.0) | 23.15 ± 5.91<br>(7.0–30.0) | 3.51 ± 2.34<br>(0.0–11.0) |

Eligible participants included individuals with a clinical diagnosis of svPPA, lvPPA, or nfvPPA based on consensus criteria^1^, with available research visits in the UCSF MAC database. For svPPA cases diagnosed with Neary-Semantic and svPPA, clinical documentation confirmed predominantly left-sided atrophy. Participants were excluded if they had not completed the picture description task, were described as having predominantly right-lateralized atrophy, were not native English speakers, or had a Clinical Dementia Rating (CDR^21^) score above 0 in the control group. PPA participants were not excluded based on MMSE or CDR in order to evaluate the transcription accuracy at a range of severities. All participants or caregivers provided informed consent following procedures aligned with the Declaration of Helsinki, and the study was approved by the UCSF Institutional Review Board.

### Disease Severity

The CDR is a widely used clinician-rated scale for disease severity that assesses cognitive and functional impairment across six domains: memory, orientation, judgment and problem-solving, community affairs, home and hobbies, and personal care^21^. Each domain is rated on a scale from 0 (no impairment) to 3 (severe impairment), and the six ratings can be summed to derive the CDR Sum of Boxes (range 0–18), which provides a fine-grained, continuous measure of cognitive and functional change. Using the same domain ratings, an established algorithm integrates this information into an overall or “global” CDR score, which ranges from 0 (cognitively normal) to 3 (severe dementia), with 0.5 indicating very mild impairment, often considered prodromal or mild cognitive impairment. The CDR Sum of Boxes and global scores are commonly used in both clinical and research settings to stage dementia severity and track progression over time.

### Speech Task

Speech was elicited using the “picnic scene” picture description task from the Western Aphasia Battery (WAB)^22^. Each participant completed the task individually in a quiet testing environment, seated across from the examiner. They were asked to describe the picture being instructed: “Tell me what you see. Talk in sentences.” Participants were given up to 3 minutes to respond, yet most participants completed their descriptions within 1–2 minutes; in our sample, the average duration was 01:43min (SD = 53sec; range = 00:30-04:26min).

### Automated Transcription and Feature Extraction

Manual and automated transcripts (fully and semi-automated) were prepared using two distinct protocols to enable consistent feature extraction. Manual transcripts were generated through SALT Services, an online transcription service. The SALT team transcribed the audio recordings using their standard conventions, which include segmenting utterances into communication units and marking pauses, speech errors, and timestamps. Examiner utterances were excluded from transcripts. These transcripts were then preprocessed to facilitate automated feature extraction: SALT codes and line breaks were removed, and the cleaned content was exported from .slt files to plain text (.txt) format.

For automated transcription, we used the OpenAI Whisper large model (large-v3) via the Python package (v20240930), an AI-powered speech recognition tool^16^. Two versions of this output were prepared for analysis. First, the fully automated Whisper transcripts were used without any modification. Second, for semi-automated transcripts, Whisper-generated transcripts were reviewed and edited using a protocol. This quality control (QC) protocol involved correcting misspellings, mishearings, homonym errors, and punctuation, as well as adjusting grammatical inconsistencies and standardizing the representation of disfluencies and speech errors according to feature extraction requirements.

Linguistic features were extracted using an open-source, Python-based pipeline developed in the lab and openly available at https://github.com/lingualab/speechmetryflow. The pipeline takes a picture description as a .txt file, and extracts around 300 features of syntax, lexico-semantics, coherence, fluency, psycholinguistics and pragmatics. From the extracted features we chose 57 known to be disrupted in PPA (e.g. Cordella et al.^23^). Features are outlined in Table 2 and described in greater detail in the supplementary materials. The pipeline was used to extract the same features from three files: preprocessed manual transcriptions, raw Whisper output, and Whisper output that had undergone QC (see Figure 1).

**Figure 1.**
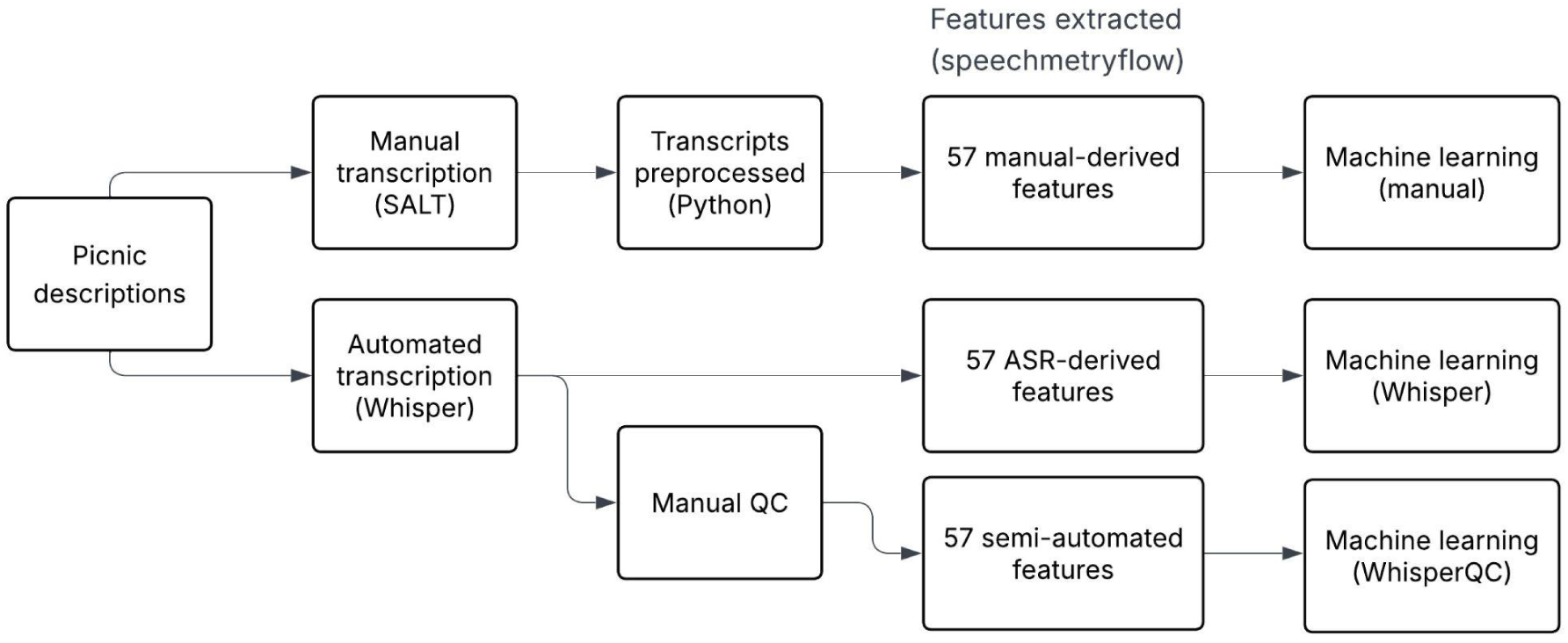
Flowchart showing transcription and feature extraction process.

**Table 2.** Features extracted from transcripts. An asterix indicates that counts were normalised by the total number of tokens in the sample.

| Domain | N features | Features |
| --- | --- | --- |
| Lexico-semantic | 4 | Indefinites (e.g. thing, stuff)*, information content units (ICUs), ICU efficiency, uncertainty words (e.g. think, maybe; list inspired by Garrard et al., 2014)*. |
| Fluency & coherence | 4 | Filled pauses (e.g. uhm, ah)*, word repetitions*, word fragments ("In her kitch kitchen")*, coherence (mean cosine distance between sentence embeddings). |
| Psycholinguistics | 20 | Mean concreteness (Brysbaert et al., 2014), frequency (SUBTLEX-US corpus; Brysbaert & New, 2009), valence (Warriner et al., 2013), familiarity and imageability (Glasgow norms; Scott et al., 2019) for all words (stop words removed), and the following POS: nouns, verbs and adjectives. |
| Lexical | 16 | Length of sample measured in tokens, parts-of-speech: nouns, pronouns, verbs, adverbs, adjectives, prepositions, determiners, and conjunctions*; plus nouns:nouns+pronouns, nouns:nouns+verbs, pronouns:nouns+pronouns, verbs:nouns+verbs; spatial and personal deictic pronouns*, moving average type-token-ratio (MATTR) (Convington & Fall, 2010). |
| Syntactic complexity | 11 | Left and right children*, subordinate clauses*, clauses per sentence*, mean length of utterance (MLU), prepositional phrases*, coordinate phrases*, incomplete sentences*. |
| Pragmatics | 2 | Formulaic expressions (e.g. as it were)*, lexical access difficulty words*. |

### Data Availability

The conditions of our ethics approval do not permit public archiving of anonymized study data. Data generated by the UCSF MAC are available upon request. Data requests can be submitted through the online UCSF MAC Resource Request form: memory.ucsf.edu/resources/data. Access will be granted to named individuals in accordance with ethical procedures governing the reuse of sensitive data. All requests will undergo a UCSF-regulated procedure, thus requiring submission of a Material Transfer Agreement, which can be found online: icd.ucsf.edu/material-transfer-and-data-agreements. No commercial use would be approved. All analysis code is available at https://github.com/lingualab/ConnectedSpeech-UCSF.

### Analysis

#### Evaluation of Transcription Accuracy

The accuracy of automated transcription was evaluated using the WER, which is the percentage of errors in an ASR-generated transcript compared to manual transcription, at the word level. The WER ranges from zero to one, with zero representing a perfect transcription and one representing 100% error. WER was calculated for both raw and QC’d transcripts, using the manual transcription as the ground truth, with the JiWER Python library (version 4.0.0). Prior to computation, both transcripts were standardized with the default preprocessing of the library. A two-way ANOVA was conducted to examine differences in WER across diagnostic groups and transcription methods (raw vs. QC), with Tukey post hoc tests ran for significant effects.

To evaluate the relationship between automated transcription accuracy and symptom severity, the Pearson correlation between WER and CDR sum was calculated using the SciPy Python library (version 1.15.3). Outlier WER scores were identified using the interquartile range (IQR). Scores falling below Q1 − 1.5×IQR or above Q3 + 1.5×IQR (where Q1 and Q3 are the first and third quartiles, and IQR = Q3 − Q1) were classified as outliers. Outliers were removed prior to the correlation analysis.

#### Reliability of Linguistic Features

Following automated extraction of the 57 linguistic features in Table 2 from all three transcriptions (manual, raw Whisper and WhisperQC), the similarity of feature values between manual and raw Whisper, and manual and WhisperQC, was evaluated using the intraclass correlation coefficient (ICC) implemented in the Pingouin Python library (version 0.5.5)^24^. The ‘ICC 3’ method was used, which is appropriate when there is no inter-rater variability, i.e., when all data are processed using the same procedure across transcription approaches, and thresholds for reliability followed Koo et al.^25^, with ICC values indicating poor (<0.5), moderate (0.5-0.75), good (0.75-0.9) or excellent (>.9) reliability. The average reliability for different linguistic domains (see Table 2) was also analysed, by calculating the mean and standard deviation of ICC values for all features belonging to a domain, before and after QC for Whisper transcripts. ICC ratings for each feature across all participants and separately for HC, svPPA, lvPPA and nfvPPA are provided in the supplementary materials, for both raw Whisper and WhisperQC transcripts.

#### Machine Learning Classification

Linguistic features extracted from the three transcript methods (manual, Whisper, WhisperQC) were used to train binary classifiers classifying HC versus svPPA, HC versus lvPPA, HC versus nfvPPA, and lvPPA versus svPPA, using the scikit-learn Python library (version 1.7.1)^26^. Fifty-seven linguistic features (see Table 2, and supplementary materials Table S1) were used as input to a linear support vector classifier (SVC), with 10-fold stratified cross-validation performed to preserve class distribution, and feature scaling conducted within each fold. Metrics included area under the curve (AUC), balanced accuracy, sensitivity, specificity, and F1-score, computed for each fold. To compare performance, we focused on the AUC and performed paired Wilcoxon signed-rank tests to assess differences between transcript methods for each classification task. The resulting *p*-values were corrected for multiple comparisons using the Benjamini–Hochberg false discovery rate (FDR) procedure to control the family-wise error rate at α = 0.05. We also investigated the impact of using only features most robust to ASR error, defined as those with an intraclass correlation rating of “good” or “excellent” in both groups being classified, for ASR-derived classifications only.

## Results

### Automated Transcription Accuracy

The mean WER for raw automated Whisper transcriptions differed across diagnostic groups, F(3, 272) = 37.10, *p* < .001. Post-hoc comparisons showed that HC had lower WERs (M = 0.13, SD = 0.18) than all three PPA variants (svPPA: M = 0.20, SD = 0.14, *p* < .001; lvPPA: M = 0.26, SD = 0.12, *p* < .001; nfvPPA: M = 0.31, SD = 0.21, *p* < .001). Among the PPA groups, svPPA had the lowest mean WER, which was lower than both lvPPA (*p* = .0015) and nfvPPA (*p* = .0007). No difference was observed between lvPPA and nfvPPA (*p* = .99). When analysing the transcribed text collapsed within each group (i.e., as opposed to per participant), there was a similar pattern (HC = 0.17, svPPA = 0.27, lvPPA = 0.30, nfvPPA = 0.36). Eight outliers with unusually high WER were identified: 2 svPPA, 2 lvPPA, and 4 nfvPPA. There was no difference in age (t(149) = –1.28, *p* = 0.239) or CDR sum score (t(149) = 0.89, *p* = 0.402) between outliers and non-outliers. Examples of outlier transcriptions using the three approaches can be found in the supplementary materials (Table S2).

Following manual QC of the raw Whisper transcriptions, the WER decreased across all groups, F(1, 272) = 40.57, *p* < .001, indicating an overall improvement in transcription accuracy (see Figure 2). The interaction between diagnosis and transcription method (raw vs QC) was not significant, F(3, 272) = 0.37, *p* = .77, indicating that group differences in WER were consistent before and after QC.

**Figure 2.**
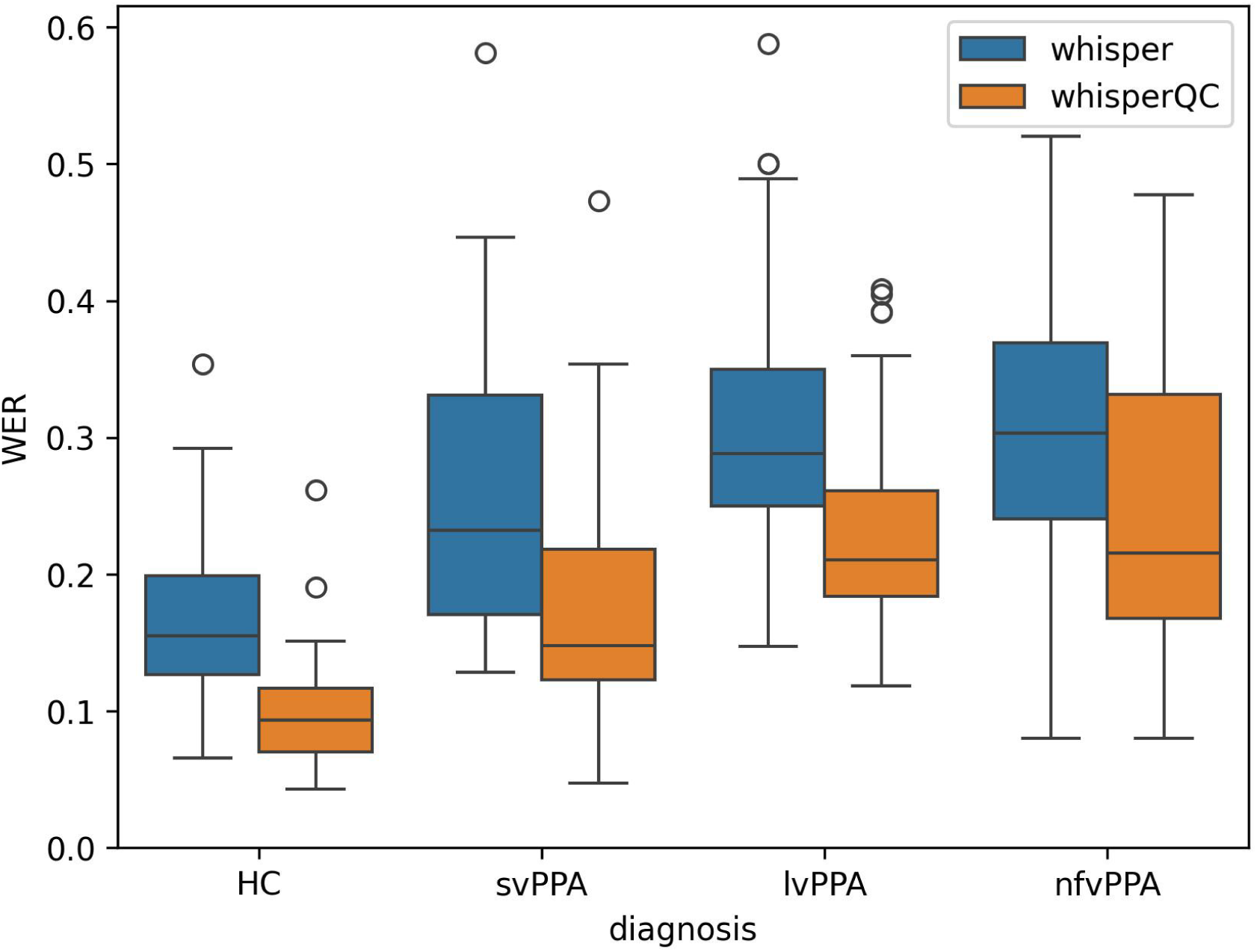
Word error rates (WER) per group for raw Whisper transcriptions in blue, and following QC, in orange. HC = healthy controls, svPPA = semantic variant primary progressive aphasia (PPA), lvPPA = logopenic variant PPA, nfvPPA = non-fluent variant PPA.

To assess whether transcription accuracy was associated with cognitive performance, Pearson correlations were computed between WER and CDR sum scores for each PPA variant (see Figure 3). There was a positive correlation between raw Whisper WERs and CDR for svPPA (*r* = 0.47, *p* = .003) and lvPPA (*r* = 0.49, *p* = .002), indicating that poorer cognitive performance was associated with greater errors in the transcription. No association was found for nfvPPA (*r* = –0.00, *p* = .99). Following QC, correlations were attenuated: svPPA remained significant (*r* = 0.34, *p* = .038), whereas the correlation for lvPPA was no longer significant (*r* = 0.27, *p* = .11), and nfvPPA continued to show no association (*r* = 0.21, *p* = .23).

**Figure 3.**
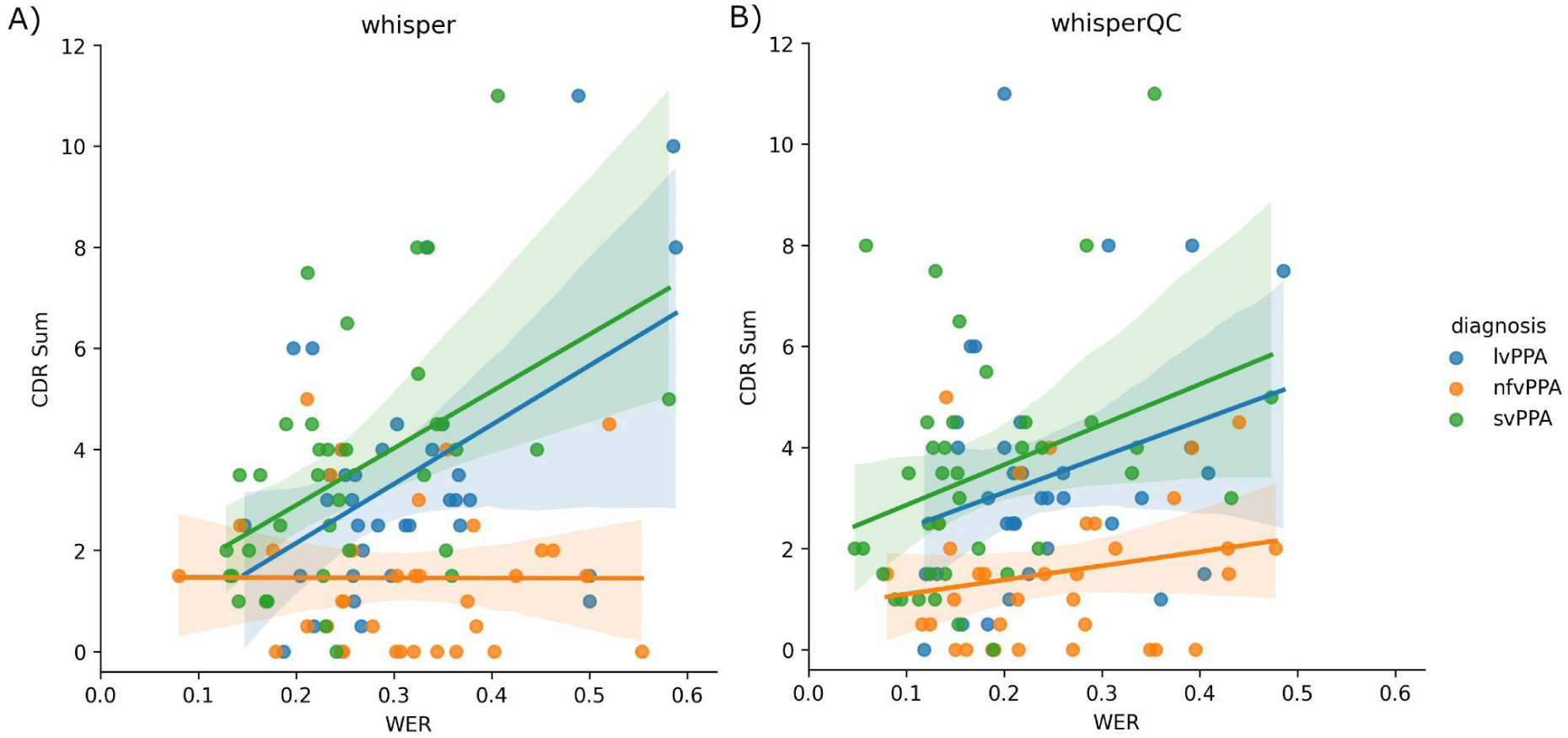
Correlation between word error rates (WER) and clinical dementia rating (CDR) sum scores, by group. A) Correlation for raw Whisper transcripts, B) correlations for WhisperQC transcripts.

### Feature Reliability

We assessed the similarity in feature values derived from Whisper generated transcripts compared to manual transcripts, by calculating the ICC of raw and QC’d Whisper outputs. Features were considered reliable if the ICC agreement was “good” (0.75-0.9) or “excellent” (>.9). Across all participants, 57.9% of features were reliable when computed from raw Whisper transcripts. Following QC this increased to 82.5%, a significant difference (χ² = 7.08, *p* = 0.008). Improvements were also observed in each subgroup: HC (71.9% to 91.2%; χ² = 5.84, *p* = 0.016), svPPA (57.9% to 91.2%; χ² = 14.98, *p* < 0.001), lvPPA (61.4% to 80.7%; χ² = 4.27, *p* = 0.039), and nfvPPA (29.8% to 66.7%; χ² = 14.05, *p* < 0.001). After correction for multiple comparisons using the Benjamini–Hochberg procedure, the improvements for svPPA and nfvPPA remained statistically significant at a *p* value threshold of 0.025.

Figure 4 shows the raw ICC values for features from raw Whisper transcripts, grouped into linguistic domains, along with thresholds for poor, moderate, good, and excellent similarity. It can be seen that features of fluency (such as filled pauses and word fragments or “false starts”) and syntactic complexity (such as clauses per sentence and mean length of sentence) generally have poor reliability, particularly for PPA variants. Fluency features also have large error bars, suggesting variability in how reliable these features are; however it should be noted that there are few features in the fluency domain. Reliability of these domains generally improves following QC (see Figure 5).

**Figure 4.**
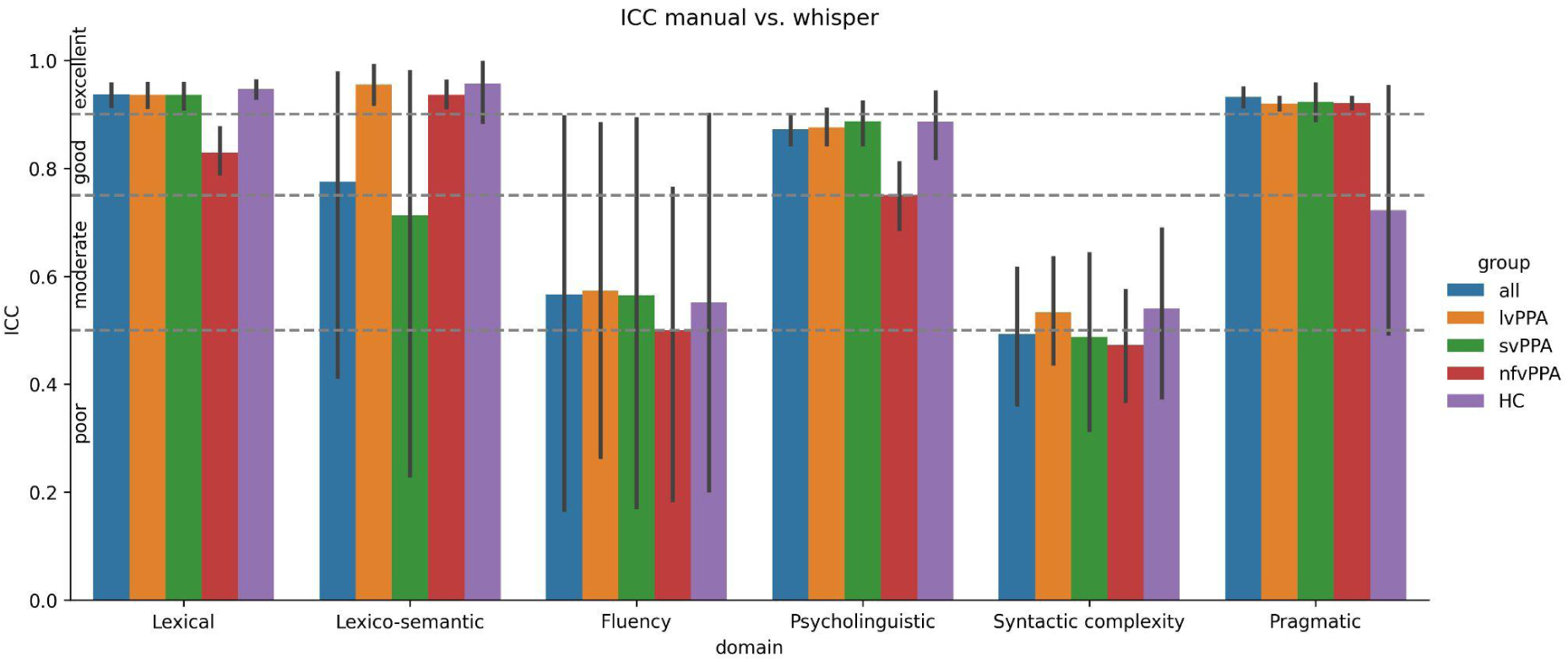
Mean intraclass correlation coefficient (ICC) for features computed from raw Whisper transcripts, within linguistic domains. Error bars are +/- standard deviation. Dotted lines indicate ICC thresholds for poor, moderate, good or excellent reliability.

**Figure 5.**
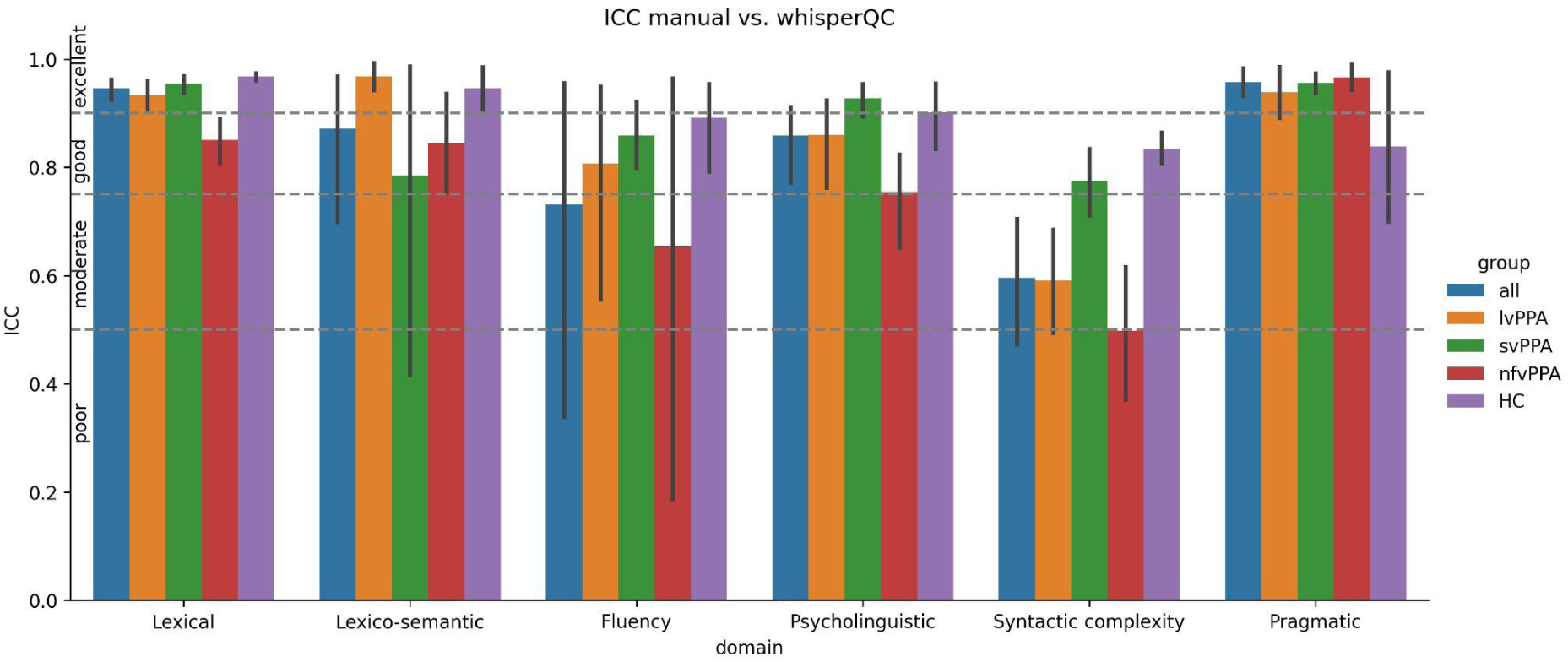
Mean intraclass correlation coefficient (ICC) for features derived from QC’d Whisper transcripts, within linguistic domains. Error bars are +/- standard deviation. Dotted lines indicate ICC thresholds for poor, moderate, good or excellent reliability.

### Classification Performance

Results of binary classifications using features derived from each transcript are shown in Table 3. Classifying HC versus svPPA, results were similar using features from all transcription methods. Features derived from raw Whisper transcripts achieved the highest performance (AUC = 0.99, compared to 0.97 for both manual and WhisperQC features). In all other classifications, features derived from Whisper outputs that had undergone manual QC achieved the highest performance. In classifying HC versus lvPPA, these features achieved an AUC of 0.98, increasing 10% from raw Whisper transcript features and 7% from manual transcript features. There was a similar pattern when classifying HC versus nfvPPA; features from WhisperQC transcripts achieved an AUC of 0.89, increasing 8% from manual transcript features, and 10% from raw Whisper transcript features; this difference was significant at *p* = 0.03, however it did not survive correction for multiple comparisons. Smaller improvements were seen when classifying lvPPA versus svPPA; WhisperQC features achieved an AUC of 0.77, increasing 3% from raw Whisper features and 2% from manual transcript features. Other performance metrics (sensitivity, specificity, balanced accuracy and f1 score) followed a similar pattern to the AUC, and can be found in the supplementary materials (Table S3).

**Table 3.** Comparison of AUCs for binary classification tasks and transcript methods using the full feature set, or with filtering to include only features robust to ASR error. Values indicate mean area under the curve (AUC) across 10-fold cross-validation (± standard deviation). The highest performance using either all features or only robust features is in bold for each method. Significant differences (uncorrected) between two transcription approaches for a classification task are indicated using *.

| Classification task | Transcript method | AUC (All features) | AUC (Robust features only) |
| --- | --- | --- | --- |
| HC-vs-svPPA | manual | 0.97 ± 0.08<br>[0.95, 1.00] | - |
|  | Whisper | **0.99 ± 0.03**<br>**[0.89, 1.00]** | **0.99 ± 0.03**<br>**[0.94, 1.00]** |
|  | WhisperQC | 0.97 ± 0.06<br>[0.86, 1.00] | 0.97 ± 0.06<br>[0.83, 1.00] |
| HC-vs-lvPPA | manual | 0.91 ± 0.11<br>[0.84, 0.96] | - |
|  | Whisper | 0.88 ± 0.16<br>[0.77, 0.98] | 0.92 ± 0.14<br>[0.77, 0.98] |
|  | WhisperQC | **0.98 ± 0.04**<br>**[0.82, 0.98]** | **0.95 ± 0.07**<br>**[0.82, 0.98]** |
| HC-vs-nfvPPA | manual | 0.81 ± 0.19<br>[0.60, 0.96] | - |
|  | Whisper | 0.79 ± 0.23*<br>[0.54, 0.86] | 0.76 ± 0.24<br>[0.56, 0.93] |
|  | WhisperQC | **0.89 ± 0.19\***<br>**[0.52, 0.93]** | **0.83 ± 0.21**<br>**[0.51, 1.00]** |
| lvPPA-vs-svPPA | manual | 0.75 ± 0.24<br>[0.41, 0.78] | - |
|  | Whisper | 0.74 ± 0.15<br>[0.38, 0.65] | 0.69 ± 0.15<br>[0.32, 0.59] |
|  | WhisperQC | **0.77 ± 0.25**<br>**[0.43, 0.84]** | **0.78 ± 0.23**<br>**[0.45, 0.81]** |

### Robust Features Only

Filtering to include only features robust to ASR error for both groups being classified, defined as “good” or “excellent” ICC reliability, reduced the set of 57 features to 44 for HC vs. svPPA, 43 for HC vs. lvPPA, 21 for HC vs. nfvPPA, and 44 for lvPPA vs. svPPA. No clear pattern of results emerged: classifying HC versus svPPA there was no difference in performance after filtering, however classifying HC versus lvPPA, filtering increased performance 4% using raw Whisper features, and decreased performance 3% using WhisperQC features. Classifying HC versus nfvPPA, filtering reduced performance for both Whisper and WhisperQC features by 3-6%, and for lvPPA versus svPPA reduced performance for Whisper transcripts by 5%, and improved performance for WhisperQC transcripts by 1% (see Table 3).

## Discussion

We compared the accuracy of gold-standard manual transcription, raw automated transcriptions using Whisper, and automated transcriptions that had undergone manual QC (WhisperQC) for transcribing speech from a large cohort of PPA variants, as well as HC. Across all classification tasks the best performance was achieved using ASR-derived features, computed from either raw Whisper transcripts or WhisperQC transcripts, suggesting that ASR transcription is acceptable, and potentially even beneficial, for detection of PPA speech errors.

Transcription errors for raw Whisper outputs, measured using the word error rate (WER), were relatively low (13% for HC, 20% for svPPA, 26% for lvPPA and 31% for nfvPPA speech), indicating good transcription accuracy. Previous research has reported higher WERs, ranging from 34% in mild aphasia^27^, up to 73.1% in svPPA^12^, and around 38.5% in post-stroke aphasia^28^. We found that the WER increased as overall disease severity, assessed using the CDR, increased, consistent with previous findings in both PPA^27^ and post-stroke aphasia^28^. The pattern of WER results by subgroup also aligns with existing literature, with lowest error rates for control samples, followed by gradually increasing error rates for svPPA, lvPPA and nfvPPA^12^. This pattern likely reflects the preserved fluency in svPPA speech, which is easier for Whisper to transcribe. Conversely, the word-finding related pauses and hesitations prevalent in lvPPA, and apraxia of speech and dysarthria seen in nfvPPA, lead to more transcription errors. There was no significant difference between WERs for lvPPA and nfvPPA, and as such findings are partly in-line with our hypothesis that nfvPPA speech would be particularly challenging for ASR.

In classification tasks, ASR-based classification of nfvPPA demonstrated lower performance overall than other variant versus HC classifications, in-line with our hypotheses, yet still achieved robust discrimination (highest AUC of 0.89 with WhisperQC-derived features) that out-performed manual-derived features (AUC of 0.81). Both HC versus svPPA and HC versus lvPPA classifications achieved very high performance, with AUCs of 0.99 (raw Whisper-derived features) and 0.98 (WhisperQC-derived features) respectively. These results indicate that ASR-derived features, particularly when manually quality-controlled, can achieve classification performance comparable to or exceeding that reported in previous studies^7,8,13,29^.

The impact of manual QC on the raw Whisper transcripts was evident across our analyses, resulting in a significant improvement for all groups in transcription accuracy (i.e., decreased WER) and reliability of features (i.e., increased number of features with “good” or “excellent” ICC reliability). Furthermore, in classifications, improvements were seen in performance between raw Whisper and WhisperQC features, with a 10% increase when classifying either lvPPA or nfvPPA versus HC. Indeed, the best performance was achieved with WhisperQC-derived features for all classification tasks except HC versus svPPA.

Including only features most robust to ASR-error did not systematically improve ASR-derived classifications (see Table 3). This lack of improvement suggests that inclusion of features that may be unreliable when using ASR does not negatively impact model performance, and in fact including poor or moderately reliable features was beneficial for some classifications. This finding reflects previous work demonstrating that feature stability was independent of feature importance in a classification model^17^. Thus, ASR-derived features may contain meaningful information about disease that is useful for classification, even when their values do not align with manual gold standard features^14,19^. In the case of svPPA, QC may have corrected ASR transcription errors that contained meaningful information, leading to a slight drop in performance compared to raw Whisper-derived features.

Our ICC analysis showed that when using ASR, features of fluency and syntax were the least reliable, also in-line with previous work showing that features depending on sentence boundaries, out of dictionary words (such as “um”), or syntactic parsing are the least stable^17^. Since nfvPPA speech is characterized by non-fluent, agrammatic speech with motor speech impairments (e.g., apraxia of speech, dysarthria), the unreliability of these features using raw Whisper outputs may have impacted classifications; manual QC of raw Whisper outputs led to improvements in both feature reliability and classification performance, with WhisperQC features out-performing manual features by 8%. Additionally, the linguistic features included in this study may not adequately capture nfvPPA motor speech deficits; for optimal classification of nfvPPA, ASR-based linguistic measures should likely be complemented with acoustic features^30^.

The lack of inclusion of acoustic features is a limitation of the current work. We also did not investigate silent pauses, since that would have necessitated a more labour-intensive semi-automated protocol in which pause information was added, but may have improved classification accuracies, since the length of silent pauses is likely informative in PPA^13,31^. Nevertheless, we achieved high AUCs on PPA versus HC classifications without silent pauses, suggesting that the additional step of analysing silent pauses may not be required. Silent pauses may be more important for distinguishing variants, in which our performance (lvPPA versus svPPA) was lower than HC classifications. A further limitation of this study is the linguistic and demographic profile of the sample, which included primarily white, highly educated, English-speaking participants. As ASR performance varies with speaker accent and linguistic background, and demographic factors such as gender^32–34^, findings may not generalise to more diverse clinical or community populations. An important next step will be to evaluate ASR for PPA across diverse linguistic groups.

Future work could also extend our analysis to other types of conversational speech. Previous research suggests that ASR is more accurate on shorter samples^35^, and at the word level on longer content words, which are common in tasks such as picture description^27^, as opposed to shorter words, which are common in free conversational speech. Since the choice of connected speech task can impact resulting linguistic features and classification performance^36^, and feature reliability also varies by task^27^, testing how an ASR pipeline impacts results for different types of connected speech in PPA would be prudent. Future studies could also look to test an automated QC step, flagging and re-processing transcripts with very few words or with a large discrepancy between audio and transcript length, reducing the time needed for manual QC.

In conclusion, we demonstrated the potential of ASR-derived features as reliable inputs for automated analysis pipelines. Fully and semi-automated Whisper-based transcripts not only approximated manual transcriptions but also supported more accurate classification. While challenges remain for more impaired or dysfluent speakers, and ASR may not be suitable for all clinical use cases, our results highlight a path toward scalable, cost-efficient tools for capturing and quantifying speech and language changes in neurodegenerative disease, particularly in the early phases of disease. Overall, our findings show that ASR can be effectively integrated into language research and clinical assessment for PPA, particularly semantic and logopenic variants, when supported by quality control procedures.

## Acknowledgments

We are grateful to the participants and their families who took part in this research.

## Conflicts of Interest

Declarations of interest: none.

## Funding Sources

This work was supported by the National Institutes of Health (NIH) under the following grants: RF1NS050915, K24DC015544, and P01AG019724 (M.L. Gorno-Tempini); R00AG066934 and R01AG091509 (J.M.J. Vonk). Natasha Clarke is funded by a Postdoctoral Fellowship from the Canadian Consortium on Neurodegeneration and Aging (CCNA) Sex & Gender Hub.

## Notes

### Competing Interest Statement

The authors have declared no competing interest.

### Author Declarations

IRB of University of California San Fransisco gave ethical approval for this work.

