## Supplementary for "Automated transcription in primary progressive aphasia: Accuracy and effects on classification"

#### Demographic distributions

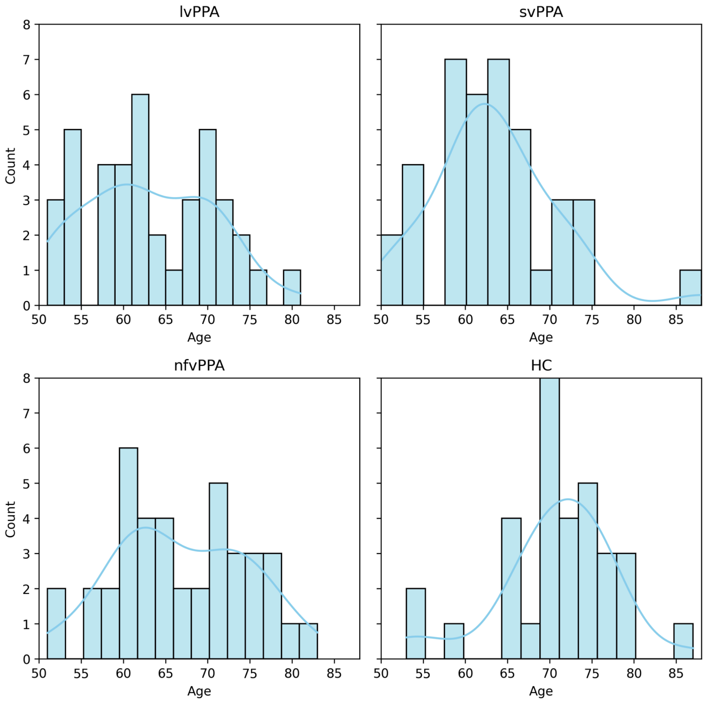

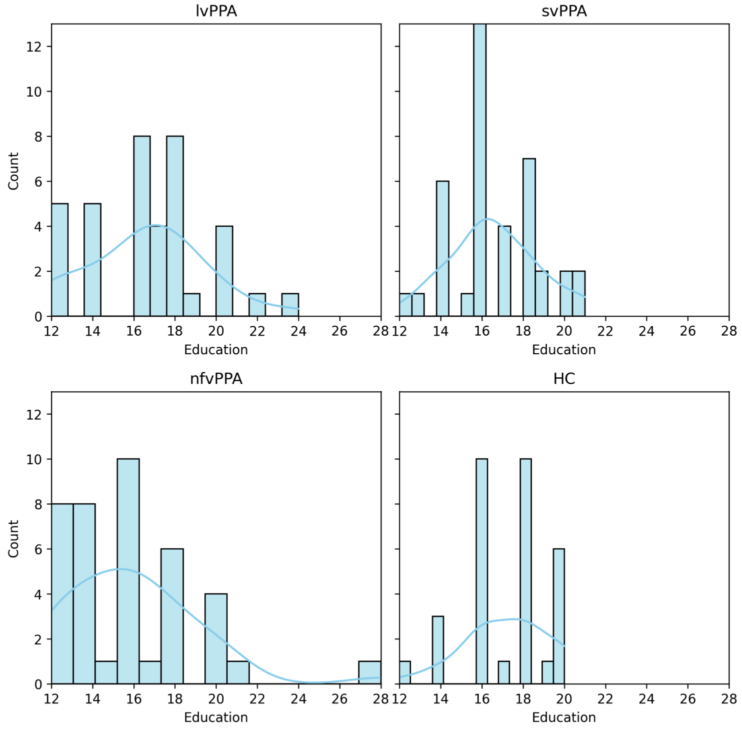

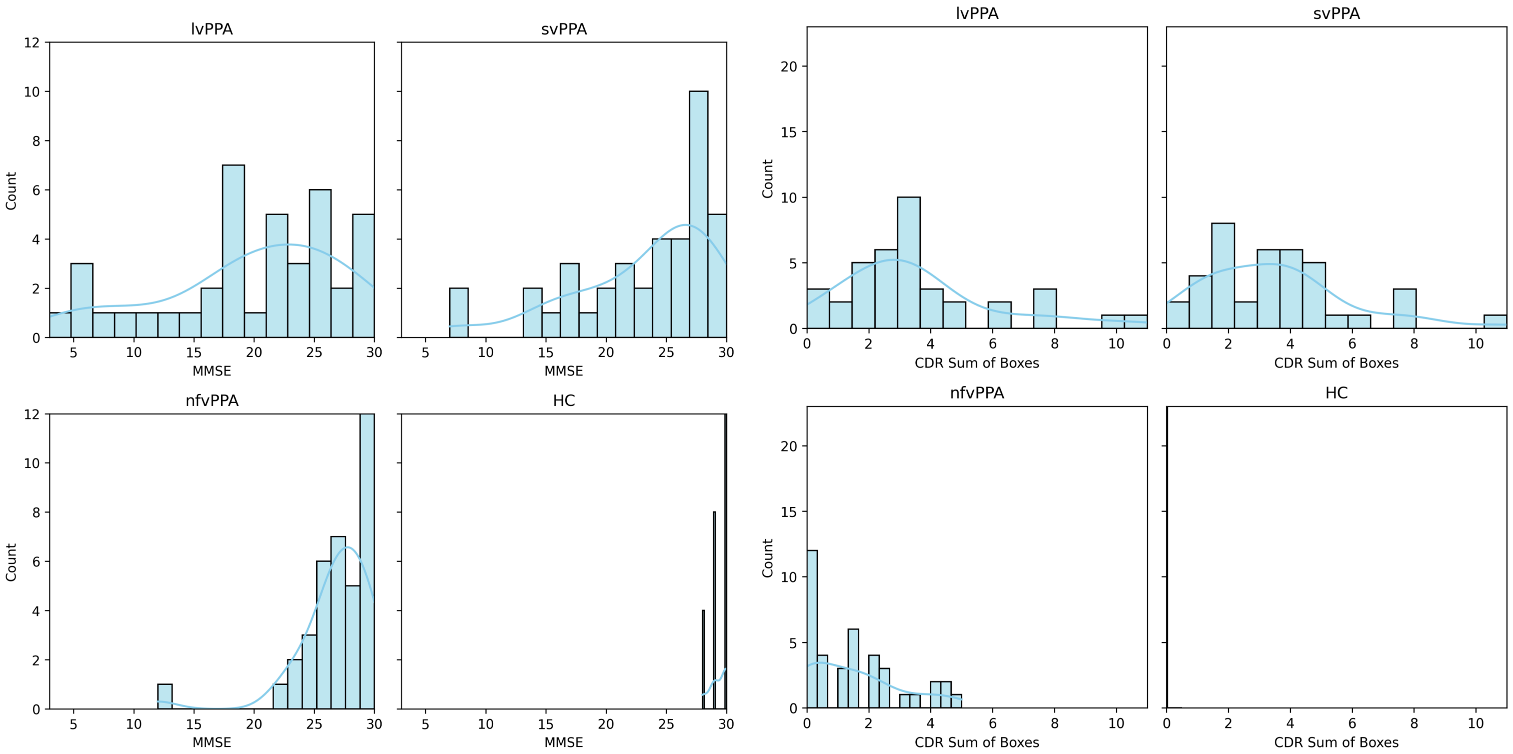

Figure S1. Histograms showing the distribution of age, years of education, Mini Mental State Examination (MMSE) scores and Clinical Dementia Rating (CDR) Sum of Boxes scores, separately for logopenic variant PPA (lvPPA), semantic variant PPA (svPPA), non-fluent variant PPA (nfvPPA) and healthy controls (HC).

### Linguistic features

| Feature | Definition | Implementation | Number of features | Notes |
|---|---|---|---|---|
| Lexico-semantic features | | | | |
| Indefinites | Non-specific words. | Total number of indefinite words* in the sample as a proportion the total number of words in the sample.<br><br>*“thing”, “stuff”, “anything”, “nothing” , “anyone”, “one”, “either”, “neither”, “everyone”, “no one”, “someone”, “anybody”, “everybody”, “nobody”, “somebody”, “another”, “the other”, “each”, “little”, “less”, “much”, “both”, “few”, “fewer”, “many”, “other”, “others”, “several” | 1 |  |
| Information Content | A measure of content using information content units (ICUs), initially defined by speech-language pathologists. ICUs include actions, subjects, places and objects within the scene. | Total number of Information Content Units (ICUs) mentioned by the speaker. | 1 | |
| Efficiency/ idea density | Ratio of the sample length to the total number of ICUs mentioned in the sample. | Total number of words in the sample divided by the total number of ICUs labelled as “TRUE”. | 1 | |

| | | | | |
|---|---|---|---|---|
| Words denoting uncertainty | Words denoting uncertainty regarding the nature of an element in the picture to be described. | Total number of uncertain words* in the sample as a proportion of total words.<br><br>*"think", "look", "like", "kind", "seem", "maybe", "can", "something" | 1 | List of words inspired by Garrard et al., 2014. |
| Fluency and coherence features | | | | |
| Local coherence | Coherence is calculated using word embeddings, which represent the meaning of a word in semantic space. The distance between these vectors indicate semantic similarity: closer embeddings are more semantically similar, and would indicate more coherent speech. | Embeddings in a sentence are averaged, and the cosine distance calculated between adjacent sentences. The mean across the sample is then calculated. | 1 | |
| Filled pauses | Pauses in the speech sample marked by "uhm" or a variation of that sound ("hmmm", "hum", "er", "ah", etc.) | Total number of occurrences of: "uhm", "hmmm", "hum", "uh", "er", "ah" in the sample. Calculated as a proportion the total number of words in the sample. | 1 | |
| Word repetitions | Words that are present more than once in the sample, either immediately or further in the sample. | Total number of words present more than once, as a proportion of the total number of words in the sample. | 1 | |
| Word | Production of only part of | Total number of word fragments in the | 1 | Example: "In her kitch |

| | | | | |
|---|---|---|---|---|
| fragments/false starts | the phonemes of a word, without sound replacements or articulation errors. May or may not be followed by the full production of the word. | sample as a proportion of the total number of words in the sample. | | kitchen" |
| Psycholinguistic features | | | | |
| Concreteness | Degree to which the concept denoted by a word refers to a perceptible entity. Concrete words are associated with the five senses, i.e. we can hear/ see/ feel/ smell or touch it, as opposed to abstract words. | Ratings taken from Brysbaert et al., 2014. The mean concreteness was calculated for all words, nouns, verbs, and adjectives in the sample. | 4 | |
| Frequency | Evaluation of the frequency with which a word is used by speakers of a language. | Ratings calculated from the SUBTLEX-US corpus (Brysbaert & New, 2009) of subtitles from film. The mean frequency was calculated for all words, nouns, verbs, and adjectives in the sample. | 4 | |
| Familiarity | Degree to which a word is known to speakers of the language. | Ratings taken from the Glasgow norms (Scott et al., 2019). The mean familiarity was obtained for all words, nouns, verbs, and adjectives in the sample. | 4 | |
| Imageability | The degree to which a mental image of the concept represented by the word can be created. | Ratings taken from the Glasgow norms (Scott et al., 2019). The mean familiarity was obtained for all words, nouns, verbs, and adjectives in the sample. | 4 | |

| | | | |
|---|---|---|---|
| Valence | Degree of agreeability of the emotions invoked by a word (ranging from unhappy to happy). | Ratings taken from Warriner et al., 2013. The mean valence was obtained for all words, nouns, verbs, and adjectives in the sample. | 4 |
| Lexical features | | | |
| Length of sample | Total number of words in a speech sample. | Total number of words (tokens) in the sample that are not punctuation markers, including filled pauses (e.g., hmmm). | 1 |
| Parts-of-speech | Parts-of-speech are defined by their grammatical usage in the context, e.g. a noun, adverb, etc. | Ratio of nouns, pronouns, verbs, adverbs, adjectives, prepositions, determiners, and conjunctions to the total number of words in the sample.<br><br>Ratios of POS using different denominators (nouns:nouns+pronouns, nouns:nouns+verbs, pronouns:nouns+pronouns, verbs:nouns+verbs) | 8<br><br>4 |
| Deictic pronouns | Pronouns that directly reference personal, or localization characteristics of the picture to be described. The specific meaning of the pronoun depends on the context in which it is used (Crystal, 2011). | Total number of occurrences of the words in the two following categories:<br><br>Spatial deictics: “this”, “that”, “here”, “there”.<br><br>Personal deictics: “he”, “she”, “her”, “herself”, “him”, “himself”,<br><br>as a proportion of the total number of words in the sample. | 2 |

| | | | | |
|---|---|---|---|---|
| Moving Average Type-Token Ratio (MATTR) | Type-Token Ratio (TTR) is a measure of word re-use, indicating lexical richness. The moving average uses a sliding window. | TTR = V/N, where V is the size of the vocabulary (different words used, or tokens), and N is the total number of words in the sample (tokens).<br>A window of size 20 words is moved through the text, the TTR calculated, and the mean of all TTRs calculated. Here, N=20. Only content words are used. | 1 | A higher MATTR indicates greater lexical diversity (Convington & Fall, 2010). |
| Syntactic complexity features | | | | |
| Left and right children | Direct dependents of a word that are connected to it by only one arc to its right or to its left in the dependency tree. | Total number of left and right children for each word in a sample, as a proportion of the total words in the sample. Calculated with the spaCy «n_left» and «n_right» commands. | 2 | https://spacy.io/usage/linguistic-features#navigating |
| Subordinate clauses | A subordinate clause is a group of words that does not express a complete thought and does not constitute a complete sentence. Complex clauses involving subordination occur when a syntactic dependent (main or not) is used as a clausal structure. | Total number of the four following universal dependencies calculated using the default spaCy dependency parser:<br><br>Clausal subjects (csubj).<br><br>Clausal complements, divided into those with mandatory control (xcomp) and those without (ccomp).<br><br>Adverbial clause modifiers (advcl).<br><br>Adnominal clause modifiers (acl),<br><br>as a proportion of the total number of | 4 | https://universaldependencies.org/u/overview/complex-syntax.html#subordination |

| | | | | |
|---|---|---|---|---|
| | | words in the sample. | | |
| Clauses per sentence | A clause is a group of words containing a verb, its subject and all modifiers. | Mean number of clauses in a sentence (calculated with the default spaCy implementation) divided by the total number of words in the sample. | 1 | |
| Mean Length of Utterance (MLU) | Mean number of words per sentence. | The mean number of words per sentence in the sample were calculated. Sentence boundaries were identified using the default spaCy dependency parser. | 1 | https://spacy.io/usage/linguistic-features#sbd |
| Prepositional phrases | Phrases that contain a preposition, its object (noun or pronoun), and any object modifier. | Total number of prepositional phrases as a proportion of the total number of words in the sample. | 1 | |
| Coordinate phrases | Phrases united by one or more coordinating conjunctions: "and", "but", "for", "nor", "or", "yet", "so". | Total number of coordinate phrases as a proportion of the total number of words in the sample. | 1 | |
| Incomplete sentences | Sentences that do not contain a minimum of one verb and its subject. | Total number of incomplete sentences in the sample as a proportion of the total number of words in the sample. | 1 | Could indicate lexical/semantic, syntactic, and/or speech planning difficulties (Boschi et al., 2017). |
| Pragmatic features | | | | |
| Formulaic expressions | Expressions with a fixed form and a non-literal meaning that have attitudinal nuances. | Total number of occurrences of the following formulaic expressions in the sample: "well", "so", "I guess", "you", "know", "as it is", "as it were", as a | 1 | |

| | | | | |
|---|---|---|---|---|
| | | proportion of the total number of words in the sample. | | |
| Word-finding difficulties | Use of words suggesting lexical access difficulties. | Total number of meta-cognitive words in the sample as a proportion of total words.<br><br>"know", "remember", "unable" | 1 | List of words inspired by Garrard et al., 2014 and Rentoumi et al., 2014. |

*Table S1. Linguistic features derived from transcribed samples.*

#### Outlier transcription examples

| Diagnosis | Manual | Whisper | WhisperQC |
|---|---|---|---|
| **nfvPPA (MMSE=22)** | So can I And the sailboat is in on the lake. And there and uh are people in the sailboat. | So, and I, and the cell boat, and I'm late, and they're, and people in the cell boat. | so, and I and hmm the sailboat, in um on lake, and there, and people in the sailboat. |
| **nfvPPA (MMSE=28)** | The um the um um um dinner um the uh um the um um parents have um the um um the um. | The dinner, the parents have the technique, | the um the um um um dinner um the um um the um um parents have um the um um the um ticnic, |
| **lvPPA (MMSE=6)** | Boy and girl and a tree. And a forest. Um and a dog. And and a boat. Oh are their car. House. What's that. A boy. | Boy and a girl and a tree and a dog and a boat. Boat. Oh, or the car. Mm-hmm. House. What's | Boy and girl and a tree and a sw- Four, four. Um, and a dog and a boat. Oh, are there cars? House. What's that? |

| | | | |
|---|---|---|---|
| | The the girl is is is um something. | that? Oh, oh. Boy, yeah. That girl is... | Oh. Oh. A boy. That girl is his um Something. |
| **lvPPA (MMSE=23)** | There's a a dog. The flag. Nice looking tree. And that's about it I guess. | There's a dog and a flag. Nice looking tree, and that's about it I guess. That's the hard part, yes. So, well now see, that's my problem now, what, that's, it's driving me nuts. | There's a dog and a flag. Nice looking tree. And that's about it, I guess. That's the hard part. So.. well now see that's my problem now...Drives me nuts |
| **svPPA (MMSE=7)** | Wow. Too much in here. Is too much. Yeah. There's somebody over there, right? Yeah? And two women, two guys. Uh there's. Nobody's. Here, this one. Here. | I don't know, it's amazing, yeah, wow, it's beautiful, you know, it's too much here, the water is too much here, yeah, oh, what is that one, I don't know, I don't know, I don't There's somebody over there, and there's two over there, two women, two guys, whatever. | I don't know, it's amazing, yeah? Wow. Just a. I know it's too much, yeah. The water is too much, yeah? Yeah. Oh, what is that one? I know, but I don't know. Huh. Somebody over there, huh? Yeah. And there's two there, two women, two guys, whatever. |

*Table S2. Excerpts from Whisper and WhisperQC transcripts identified as having unusually higher error rates (outliers) when compared to Manual transcripts, using the word error rate.*

### ICC ratings for each linguistic variable, per group

#### Raw Whisper

| feature | all | HC | svPPA | lvPPA | nfvPPA |
|---|---|---|---|---|---|
| Adjective_percentage | good | excellent | excellent | excellent | moderate |
| Adverb_percentage | excellent | good | excellent | excellent | good |
| Coherence_local | poor | poor | moderate | poor | poor |
| Complements_Clausal_Controlled_relative | good | excellent | good | good | moderate |
| Complements_Clausal_Non_Controlled_relative | moderate | poor | poor | poor | moderate |
| Concreteness_average_adjectives | good | good | good | moderate | moderate |
| Concreteness_average_words | excellent | excellent | excellent | excellent | good |
| Concreteness_average_nouns | excellent | excellent | excellent | excellent | poor |
| Concreteness_average_verbs | excellent | excellent | excellent | excellent | excellent |
| Coordinating_conjunctions_percentage | excellent | excellent | excellent | excellent | excellent |
| Determiner_percentage | excellent | excellent | excellent | good | moderate |
| Efficiency_ICU | poor | excellent | poor | excellent | excellent |
| Familiarity_average_adjectives | good | good | good | good | good |
| Familiarity_average_words | good | excellent | good | good | good |
| Familiarity_average_nouns | good | excellent | excellent | good | good |
| Familiarity_average_verbs | good | good | good | good | good |
| Frequency_average_adjectives | good | good | good | good | good |
| Frequency_average_words | excellent | good | excellent | good | good |
| Frequency_average_nouns | moderate | moderate | excellent | excellent | poor |
| Frequency_average_verbs | excellent | poor | excellent | excellent | excellent |
| Frequency_lexical_access_difficulty_words_relative | excellent | poor | excellent | excellent | excellent |
| Frequency_formulaic_expressions_relative | excellent | excellent | good | excellent | excellent |

| | | | | | |
|---|---|---|---|---|---|
| **Frequency_uncertainty_words_relative** | excellent | excellent | excellent | excellent | excellent |
| **Frequency_coordinated_sentences_relative** | poor | poor | moderate | moderate | poor |
| **Imageability_average_adjectives** | good | good | good | good | good |
| **Imageability_average_mots** | excellent | excellent | excellent | good | good |
| **Imageability_average_noms** | good | excellent | good | excellent | moderate |
| **Imageability_average_verbes** | good | excellent | good | good | moderate |
| **Mean_sentence_length** | poor | poor | poor | poor | poor |
| **MATTR_25** | excellent | excellent | good | good | excellent |
| **Adnomial_clausal_modifiers_relative** | moderate | good | good | poor | poor |
| **Adverbial_clausal_modifiers_relative** | moderate | moderate | poor | moderate | good |
| **Average_children_right** | poor | good | poor | poor | poor |
| **Average_children_left** | moderate | moderate | moderate | moderate | moderate |
| **N_clauses_per_sentence** | poor | poor | poor | poor | poor |
| **Noun_percentage** | excellent | excellent | excellent | good | good |
| **N_ICU_TRUE** | excellent | excellent | excellent | excellent | excellent |
| **N_fragments** | good | good | good | excellent | moderate |
| **N_words** | excellent | excellent | excellent | excellent | excellent |
| **N_filled_pauses** | poor | poor | poor | poor | poor |
| **N_incomplete_sentences_relative** | moderate | moderate | moderate | good | poor |
| **N_prepositional_phrases_relative** | moderate | poor | moderate | moderate | poor |
| **N_pronouns_deictic_personal** | excellent | excellent | excellent | excellent | excellent |
| **N_pronouns_deictic_spatial** | excellent | excellent | excellent | excellent | good |
| **N_repetitions_words** | good | excellent | good | good | good |
| **Nouns/(Nouns+Pronouns)** | excellent | excellent | excellent | excellent | good |
| **Nouns/(Nouns+Verbs)** | excellent | excellent | excellent | excellent | good |
| **Preposition_percentage** | excellent | excellent | excellent | excellent | good |
| **Pronoun_percentage** | excellent | excellent | excellent | good | good |

| | | | | | |
|---|---|---|---|---|---|
| **Pronouns/(Nouns+Pronouns)** | excellent | excellent | excellent | excellent | good |
| **Ratio_indefinite_terms** | excellent | good | excellent | excellent | excellent |
| **Valence_average_adjectives** | good | excellent | good | good | good |
| **Valence_average_words** | excellent | excellent | excellent | excellent | moderate |
| **Valence_average_nouns** | excellent | excellent | excellent | excellent | good |
| **Valence_average_verbs** | good | excellent | moderate | moderate | good |
| **Verb_percentage** | good | excellent | good | excellent | moderate |
| **Verbs/(Nouns+Verbs)** | excellent | excellent | excellent | excellent | good |

#### Whisper QC

| feature | all | HC | svPPA | lvPPA | nfvPPA |
|---|---|---|---|---|---|
| **Adjective_percentage** | excellent | excellent | excellent | excellent | good |
| **Adverb_percentage** | excellent | excellent | excellent | excellent | good |
| **Coherence_local** | poor | moderate | good | poor | poor |
| **Complements_Clausal_Controlled_relative** | good | excellent | good | moderate | moderate |
| **Complements_Clausal_Non_Controlled_relative** | moderate | good | moderate | poor | moderate |
| **Concreteness_average_adjectives** | good | excellent | excellent | good | moderate |
| **Concreteness_average_words** | excellent | excellent | excellent | excellent | good |
| **Concreteness_average_nouns** | good | excellent | good | excellent | moderate |
| **Concreteness_average_verbs** | good | excellent | excellent | good | excellent |
| **Coordinating_conjunctions_percentage** | excellent | excellent | excellent | excellent | excellent |
| **Determiner_percentage** | good | excellent | excellent | good | moderate |
| **Efficiency_ICU** | moderate | excellent | poor | excellent | moderate |
| **Familiarity_average_adjectives** | excellent | excellent | excellent | good | good |

| | | | | | |
|---|---|---|---|---|---|
| **Familiarity_average_words** | excellent | excellent | good | excellent | good |
| **Familiarity_average_nouns** | excellent | excellent | excellent | excellent | moderate |
| **Familiarity_average_verbs** | good | good | good | excellent | good |
| **Frequency_average_adjectives** | good | good | excellent | moderate | good |
| **Frequency_average_words** | good | moderate | good | good | moderate |
| **Frequency_average_nouns** | poor | moderate | excellent | poor | poor |
| **Frequency_average_verbs** | excellent | moderate | excellent | excellent | excellent |
| **Frequency_lexical_access_difficulty_words_relative** | excellent | moderate | excellent | excellent | excellent |
| **Frequency_formulaic_expressions_relative** | excellent | excellent | excellent | good | excellent |
| **Frequency_uncertainty_words_relative** | excellent | excellent | excellent | excellent | excellent |
| **Frequency_coordinated_sentences_relative** | moderate | good | good | moderate | poor |
| **Imageability_average_adjectives** | good | excellent | excellent | good | good |
| **Imageability_average_mots** | excellent | excellent | excellent | excellent | excellent |
| **Imageability_average_noms** | excellent | excellent | good | excellent | good |
| **Imageability_average_verbes** | good | good | excellent | good | good |
| **Mean_sentence_length** | poor | good | good | poor | poor |
| **MATTR_25** | excellent | excellent | good | good | excellent |
| **Adnomial_clausal_modifiers_relative** | moderate | good | good | moderate | poor |
| **Adverbial_clausal_modifiers_relative** | good | good | good | good | good |
| **Average_children_right** | poor | moderate | moderate | poor | poor |
| **Average_children_left** | moderate | good | moderate | moderate | moderate |
| **N_clauses_per_sentence** | poor | good | moderate | poor | poor |
| **Noun_percentage** | excellent | excellent | excellent | excellent | good |
| **N_ICU_TRUE** | excellent | good | excellent | excellent | excellent |
| **N_fragments** | good | excellent | good | good | good |
| **N_words** | excellent | excellent | excellent | excellent | excellent |
| **N_filled_pauses** | excellent | excellent | excellent | excellent | excellent |

| | | | | | |
|---|---|---|---|---|---|
| N_incomplete_sentences_relative | good | good | excellent | good | moderate |
| N_prepositional_phrases_relative | good | good | good | good | moderate |
| N_pronouns_deictic_personal | excellent | excellent | excellent | excellent | excellent |
| N_pronouns_deictic_spatial | excellent | excellent | excellent | excellent | good |
| N_repetitions_words | excellent | excellent | excellent | excellent | excellent |
| Nouns/(Nouns+Pronouns) | excellent | excellent | excellent | excellent | good |
| Nouns/(Nouns+Verbs) | excellent | excellent | excellent | excellent | good |
| Preposition_percentage | excellent | excellent | excellent | excellent | good |
| Pronoun_percentage | excellent | excellent | excellent | excellent | good |
| Pronouns/(Nouns+Pronouns) | excellent | excellent | excellent | excellent | good |
| Ratio_indefinite_terms | excellent | excellent | excellent | excellent | good |
| Valence_average_adjectives | excellent | excellent | excellent | excellent | good |
| Valence_average_words | excellent | excellent | excellent | excellent | good |
| Valence_average_nouns | excellent | excellent | excellent | excellent | good |
| Valence_average_verbs | good | excellent | good | good | good |
| Verb_percentage | good | excellent | good | good | moderate |
| Verbs/(Nouns+Verbs) | excellent | excellent | excellent | excellent | good |

### Classification performance

| Task | Transcript type | AUC | Balanced accuracy | F1 | Sensitivity | Specificity |
|---|---|---|---|---|---|---|
| HC-vs-svPPA | manual | 0.97 ± 0.08 [0.95, 1.00] | 0.91 ± 0.10 [0.80, 1.00] | 0.92 ± 0.09 [0.77, 1.00] | 0.93 ± 0.12 [0.62, 1.00] | 0.90 ± 0.16 [0.86, 1.00] |

| | | | | | | |
|---|---|---|---|---|---|---|
| | Whisper | **0.99 ± 0.03 [0.89, 1.00]** | **0.94 ± 0.10 [0.77, 1.00]** | **0.95 ± 0.09 [0.72, 1.00]** | **0.95 ± 0.11 [0.56, 1.00]** | **0.93 ± 0.14 [0.86, 1.00]** |
| | WhisperCQ | 0.97 ± 0.06 [0.86, 1.00] | 0.86 ± 0.07 [0.73, 1.00] | 0.87 ± 0.06 [0.71, 1.00] | 0.88 ± 0.13 [0.62, 1.00] | 0.84 ± 0.17 [0.71, 1.00] |
| HC-vs-lvPPA | manual | 0.91 ± 0.11 [0.84, 0.96] | 0.81 ± 0.11 [0.69, 0.94] | 0.80 ± 0.13 [0.72, 0.93] | 0.78 ± 0.22 [0.75, 0.88] | 0.84 ± 0.17 [0.57, 1.00] |
| | Whisper | 0.88 ± 0.16 [0.77, 0.98] | 0.81 ± 0.19 [0.69, 0.94] | 0.84 ± 0.15 [0.67, 0.93] | 0.85 ± 0.17 [0.56, 0.88] | 0.77 ± 0.27 [0.57, 1.00] |
| | WhisperCQ | **0.98 ± 0.04 [0.82, 0.98]** | **0.89 ± 0.11 [0.66, 0.93]** | **0.90 ± 0.10 [0.62, 0.94]** | **0.88 ± 0.13 [0.50, 1.00]** | **0.91 ± 0.15 [0.57, 1.00]** |
| HC-vs-nfvPPA | manual | 0.81 ± 0.19 [0.60, 0.96] | 0.77 ± 0.16 [0.55, 0.88] | 0.75 ± 0.20 [0.48, 0.88] | 0.70 ± 0.26 [0.38, 0.88] | **0.83 ± 0.18 [0.57, 1.00]** |
| | Whisper | 0.79 ± 0.23 [0.54, 0.86] | 0.76 ± 0.18 [0.47, 0.87] | 0.76 ± 0.21 [0.44, 0.88] | 0.72 ± 0.25 [0.38, 0.88] | 0.80 ± 0.17 [0.43, 0.86] |
| | WhisperCQ | **0.89 ± 0.19 [0.52, 0.93]** | **0.85 ± 0.17 [0.47, 0.93]** | **0.87 ± 0.15 [0.48, 0.93]** | **0.88 ± 0.18 [0.43, 0.94]** | **0.83 ± 0.24 [0.43, 1.00]** |
| lvPPA-vs-svPPA | manual | 0.75 ± 0.24 [0.41, 0.78] | 0.68 ± 0.21 [0.44, 0.75] | 0.67 ± 0.22 [0.40, 0.76] | 0.69 ± 0.26 [0.38, 0.88] | 0.68 ± 0.24 [0.25, 0.88] |
| | Whisper | 0.74 ± 0.15 [0.38, 0.65] | 0.68 ± 0.12 [0.38, 0.66] | 0.68 ± 0.12 [0.35, 0.70] | 0.69 ± 0.16 [0.38, 0.82] | 0.68 ± 0.17 [0.25, 0.62] |
| | WhisperCQ | **0.77 ± 0.25 [0.43, 0.84]** | **0.71 ± 0.19 [0.44, 0.81]** | **0.70 ± 0.21 [0.48, 0.81]** | **0.70 ± 0.26 [0.50, 1.00]** | **0.72 ± 0.22 [0.25, 0.75]** |

*Table S3. Performance of transcript-derived linguistic features (full set of 57 features) across binary classification tasks (support vector classifier), for five metrics. AUC = area under the curve. For each task, the best performing result for each metric is in bold.*
